# Increased burden of influenza A/H1N1pdm09 in older adults following the COVID-19 pandemic

**DOI:** 10.64898/2026.05.20.26353664

**Authors:** Simon P. J. de Jong, Colin A. Russell

## Abstract

Of the two influenza A virus (IAV) subtypes circulating endemically in humans, A/H3N2 and A/H1N1pdm09, A/H3N2 has historically been the dominant driver of disease burden in older adults. Based on an analysis of publicly available global surveillance data from 2015 to 2025 (>300,000 subtyped, age-stratified infections), we report a substantially increased contribution of A/H1N1pdm09 to influenza morbidity in older adults since approximately 2022. Birth cohort-stratified analyses suggest elevated A/H1N1pdm09 burden among individuals born before 1955-1959, consistent with erosion of pre-existing immunity originally generated by exposure to historical A/H1N1 strains. Pooled estimates across datasets and analytical approaches indicate the increase in A/H1N1pdm09 burden rises with earlier birth year, ranging from 1.22-fold (95% CI 1.08-1.37) for the 1955-1959 birth cohort to 3.10-fold (95% CI 2.58-3.72) for the 1930-1934 cohort. These findings point to a substantial rise in the overall influenza burden among the most vulnerable age groups, with implications for vaccine policy, clinical management, and public health planning.

## Main text

Older adults are at markedly elevated risk of severe outcomes from influenza. Of the two influenza A virus (IAV) subtypes currently circulating in humans, A/H3N2 and A/H1N1pdm09, A/H3N2 has historically disproportionately contributed to influenza burden in older adults^1,2^. However, emerging evidence suggests possible shifts in this epidemiological landscape. A recent US-based study reported increasing contributions of A/H1N1pdm09 virus infections to influenza hospitalizations among older adults since the COVID-19 pandemic^3^. By contrast, in Norway a lower contribution of older adults to A/H3N2 infections was observed in this period, with limited change for A/H1N1pdm09^4^. Shifts in influenza epidemiology among older adults could translate into substantial public health consequences owing to their elevated risk of severe outcomes^5,6^; hence, a deeper understanding of these trends is needed. Here, we synthesize published global age- and subtype-resolved influenza surveillance data to interrogate changes in age-specific influenza disease burden among older adults, contrasting patterns before (2015-2020) and following (2023-2025) the suppression of influenza virus circulation during the COVID-19 pandemic.

First, we compiled a set of publicly available age- and subtype-resolved influenza surveillance data, each covering the periods both before and after the COVID-19 pandemic: hospitalization records from Brazil (SIVEP-Gripe; *n* = 66,219), sequence metadata from GISAID^7^ (*n* = 229,785), laboratory-confirmed cases from Australia (NNDSS; *n* = 93,850), and a set of recently published subtype-specific relative hospitalization rates estimated for the US (FluSurv-Net)^3^. For the 2015-2020 and 2023-2025 periods, we then calculated odds ratios reflecting the birth cohort-specific likelihood a subtyped IAV sample is A/H1N1pdm09 (i.e., rather than A/H3N2); we treat the within-pandemic periods (2021-2022) separately below. A key advantage of this subtype odds ratio is its robustness to changes in reporting and healthcare-seeking behavior, due to the subtypes’ similar clinical presentation. We used birth cohort, rather than age, given the known effect of early-life influenza virus exposures on subsequent infection risk^2,8–10^. Throughout, we used the 1965-1974 birth cohort as the reference against which changes are estimated, but we show below that conclusions are robust to choice of reference cohort.

Across datasets, subtype odds ratios were similar across the 2015-2020 and 2023-2025 periods for birth cohorts born between approximately 1960 and 2000 (Fig. 1a). In contrast, they diverged sharply for cohorts born before approximately 1955-1959 (Fig. 1a). In the 2015-2020 period, A/H1N1pdm09 odds declined progressively with earlier birth year before ∼1960, such that with decreasing birth year, infecting IAV subtypes were increasingly biased toward A/H3N2 (Fig. 1a). In the 2023-2025 period, the increasing bias toward A/H3N2 (i.e., a lower subtype odds ratio) with earlier birth year was strongly attenuated (Fig. 1a). Changes in subtype odds ratios were consistent across countries in the GISAID data (Fig. 1b). The consistency of these patterns across geographically heterogeneous datasets strongly suggests changes in underlying birth cohort- and subtype-specific influenza burden, with increasing prominence among progressively earlier birth cohorts beginning around 1955-1959. This timing is notable given the role of pre-existing immunity to historical A/H1N1 viruses (which circulated up to the 1957/1958 A/H2N2 pandemic) in reducing susceptibility to influenza A/H1N1pdm09^2,11–14^. A temporally resolved analysis of the global GISAID data, including the pandemic phase (2021-2022), places the shift in approximately 2022 (Fig. 1c), which supports defining the post-pandemic period as 2023-2025 to maximally separate pre- and post-shift epidemiological regimes.

**Figure 1.**
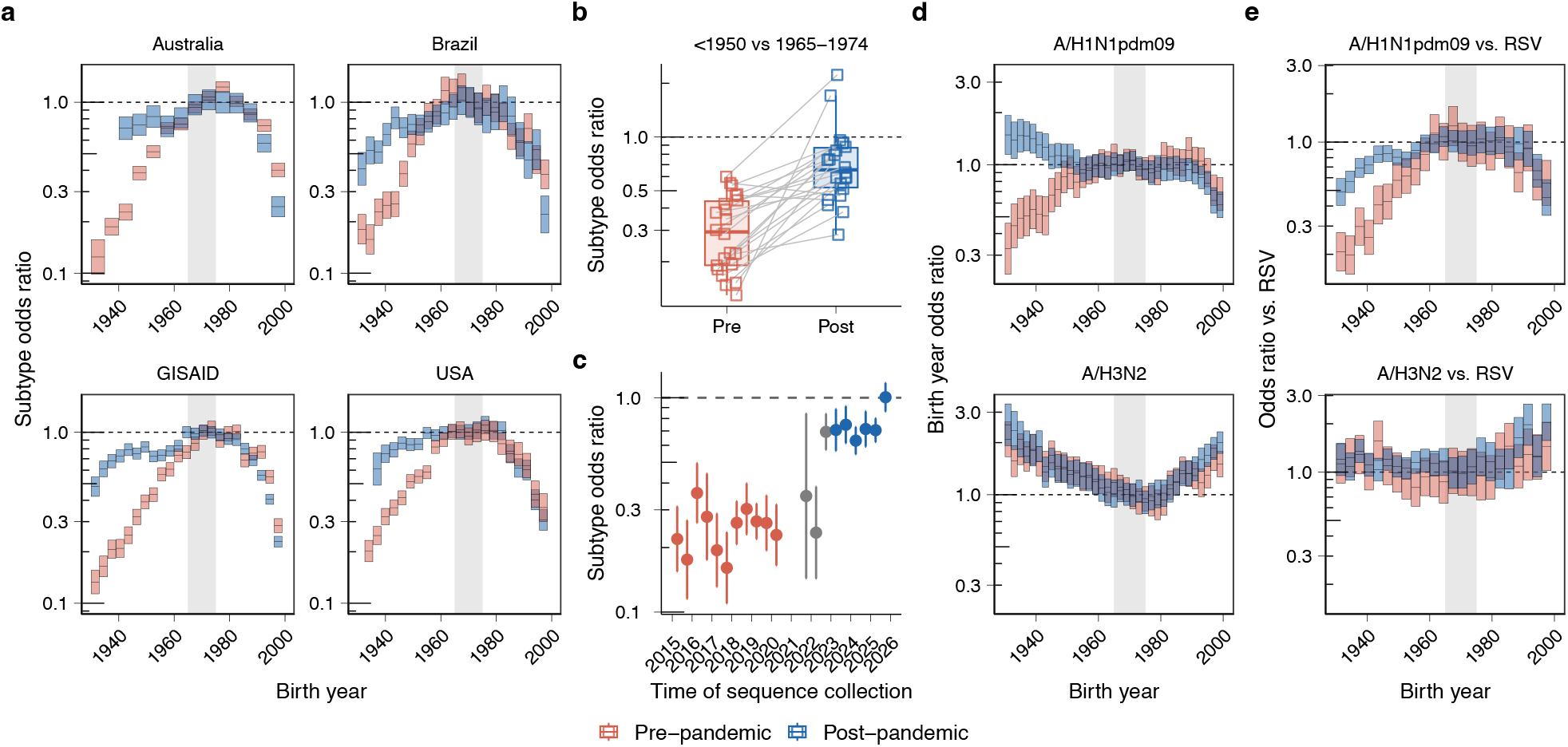
Increased burden of influenza A/H1N1pdm09 in early birth cohorts. **a** Odds a subtyped IAV sample is A/H1N1pdm09 by birth cohort, relative to the 1965-1974 reference birth cohort, by dataset, for 2015-2020 vs 2023-2025. Rectangle width and height here and in **d** and **e** correspond to age bin span and 95% CI respectively; grey vertical bars indicate the reference birth cohort. **b** Subtype odds ratio among individuals born <1950 compared to those born 1965-1974, by country in the GISAID dataset among countries with >10 sequences in both time periods (i.e., 2015-2020 and 2023-2025) and both birth cohorts (*n* = 22). **c** Subtype odds ratio (<1950 vs 1965-1974) by six-month bin of GISAID sample collection. Grey points correspond to time bins outside the defined pre-(2015-2020) and post-pandemic (2023-2025) periods. Error bars are 95% CIs. **d** Odds that the originating host of a A/H1N1pdm09 (top) or A/H3N2 (bottom) sequence was born in a year on the *x*-axis, relative to birth in 1965-1974. Estimates are from a negative binomial mixed-effects generalized linear model fitted to the GISAID data. **e** Odds a patient is positive for A/H1N1pdm09 (top) or A/H3N2 (bottom) relative to RSV, by birth cohort, relative to the 1965-1974 reference cohort, in the Brazilian data by time period.

A key question is whether the shift in subtype odds ratios among early birth cohorts reflects an increase in A/H1N1pdm09 burden, a decrease in A/H3N2 burden, or potentially both. We used a mixed-effects generalized linear model to investigate whether the birth year distribution of sample hosts in the GISAID dataset had changed since the COVID-19 pandemic for either subtype, adjusting for mortality and incorporating heterogeneity across countries. We found that in the pre-pandemic period, earlier birth cohorts were strongly underrepresented among the originating hosts of A/H1N1pdm09 sequences relative to individuals born 1965-1974, with this underrepresentation increasing with earlier birth year (Fig. 1d). Following the pandemic, in contrast, the underrepresentation of early birth cohorts was absent (Fig. 1d). Birth year distributions of A/H3N2 sequence hosts were highly similar pre-and post-pandemic (Fig. 1d). While results from any single country could be affected by changes in reporting or sequencing practice, the consistency of the pattern across the >100 countries contributing to GISAID strongly suggests an increase in A/H1N1pdm09 burden among early birth cohorts.

To further control for changes in testing and healthcare-seeking behavior, we compared A/H1N1pdm09 infection frequency in early birth cohorts against respiratory syncytial virus (RSV) and human parainfluenza virus (HPIV) infections in the Brazilian hospitalization data, reasoning that a genuine increase in A/H1N1pdm09 incidence should be detectable against these non-influenza comparators. Pre-pandemic, A/H1N1pdm09 was strongly underrepresented compared to RSV in early birth cohorts relative to the 1965-1974 reference; post-pandemic, this underrepresentation was substantially attenuated (Fig. 1e, top). Birth cohort distributions of A/H3N2 relative to RSV were highly similar pre- and post-pandemic (Fig. 1e, bottom), and similar patterns were observed for HPIV (Extended Data Fig. 1). The absence of clear change for A/H3N2, but strong evidence of change for A/H1N1pdm09, across both the RSV/HPIV control analyses and the GISAID birth year enrichment analysis above supports an increase in older adult A/H1N1pdm09 burden driving changes in subtype odds ratios seen in Fig. 1a.

Collectively, our analyses indicate that influenza burden due to A/H1N1pdm09 viruses has substantially increased among older birth cohorts. Whereas early birth cohorts were strongly underrepresented among A/H1N1pdm09 cases in the pre-pandemic period, this underrepresentation is markedly attenuated post-pandemic. To quantify the magnitude of pre- to post-pandemic increases in A/H1N1pdm09 burden, we derived birth cohort-specific fold increases from the above analyses; specifically, from subtype odds ratios across the US, Australian, Brazilian, and GISAID datasets, birth year enrichment in the GISAID dataset, and comparisons against RSV and HPIV the Brazilian dataset. Then, we combined all seven in a multilevel random-effects pooled analysis, drawing on the breadth of data sources and analytical approaches to yield a robust overall estimate. Estimated fold increases ranged from 1.22-fold (95% CI 1.08-1.37) for the 1955-1959 cohort to 3.10-fold (95% CI 2.58-3.72) for the 1930-1934 cohort (Fig. 2) and were robust to choice of reference birth cohort (Extended Data Table 1). Individual estimates were highly consistent across datasets and analytical approaches (Fig. 2).

**Figure 2.**
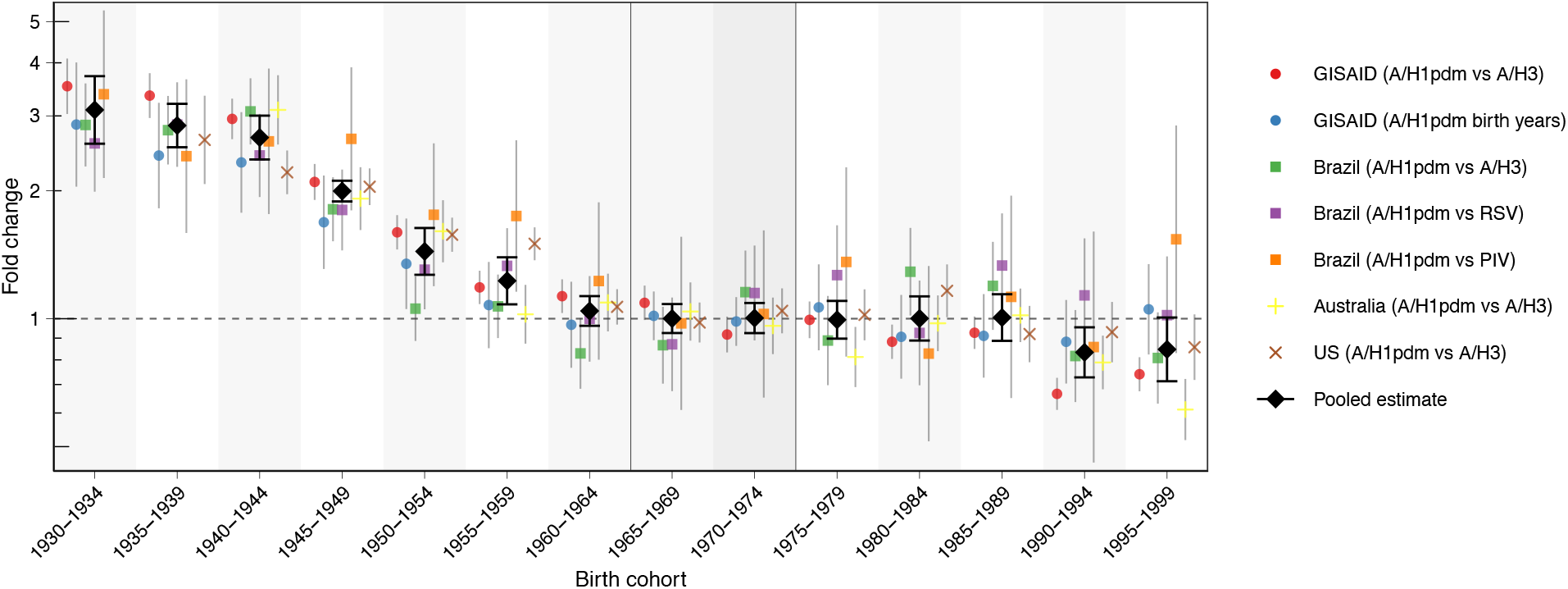
Fold change in A/H1N1pdm09 burden by birth cohort. Points and lines reflect estimates and 95% CIs of the fold change in A/H1N1pdm09 burden from 2015-2020 to 2023-2025 by birth cohort, across datasets and analytical approaches. Black diamonds reflect pooled estimates from a multilevel random-effects pooled analysis. Vertical grey lines denote the 1965-1974 reference cohort.

Increased A/H1N1pdm09 burden begins with cohorts born around 1955–1959 and increases with progressively earlier birth year (Fig. 2). This corresponds to birth cohorts alive prior to the 1957–1958 A/H2N2 pandemic, which displaced the previously circulating A/H1N1 lineage. Pre-existing immunity to historical A/H1N1 viruses, acquired through early-life exposure, has been associated with reduced susceptibility to A/H1N1pdm09 disease, attributed to cross-reactive antibody responses^10–12,15^. We thus hypothesize that contemporary A/H1N1pdm09 viruses have evolved antigenically away from the epitopes targeted by pre-existing cross-reactive immunity, eroding the protection these cohorts had previously enjoyed.

The effect boundary around 1955–1959 implicates not only cohorts with early-life exposure to 1918-lineage H1N1 viruses^3^, but also those exposed to the antigenically distinct A-prime H1N1 strains that emerged in the 1940s and circulated until 1957. The lack of signal of any analogous shift among individuals born 1977–2009 (who experienced childhood exposure to A-prime-like H1N1 viruses following their reintroduction) may help constrain which specific cross-reactive responses are implicated. The pattern of larger fold increases with earlier birth cohort (Fig. 2) may reflect the stronger pre-existing immunity in these cohorts (i.e., a larger baseline level of protection available to be eroded) potentially driven by more pronounced immunological imprinting due to a greater number of cumulative early-life exposures to historical A/H1N1 viruses and/or greater antigenic similarity of A/H1N1pdm09 to 1918-like A/H1N1 viruses^2,11^. The post-pandemic epidemiological shift coincides with the global emergence of the 6B.1A.5a.2a A/H1N1pdm09 clade (Extended Data Fig. 2); whether antigenic evolution associated with this clade drove loss of cross-protection warrants investigation.

The apparent increases in A/H1N1pdm09 risk are greatest in the earliest birth cohorts (Fig. 2). This could have substantial clinical and public health implications, as risk of severe influenza disease and mortality increases strongly with age^5,6^. As such, our results have implications for healthcare capacity planning, with likely substantially elevated acute and post-acute healthcare demand due to influenza compared to a counterfactual in which no immune erosion had occurred. The severity of the 2024/2025 US influenza season potentially reflects the erosion of protection in early birth cohorts^3^, with adults aged 75 and over experiencing the highest rates of A/H1N1pdm09-associated hospitalization since the 2009 pandemic^16^.

Vaccines have demonstrated effectiveness against A/H1N1pdm09 among adults aged 65 and over in the post-pandemic period^17,18^, but new targeted analyses in the earliest birth cohorts, for which estimated increase in burden is greatest (Fig. 2), would be valuable. A potential concern is that individuals whose immunity was shaped predominantly by early-life responses to historical A/H1N1 epitopes may, through repeated boosting of those same responses, lack immunity to epitopes on strains circulating since 2022, leaving them with little cross-reactive protection to recall upon vaccination or infection. These uncertainties notwithstanding, the elevated A/H1N1pdm09 risk suggested here further strengthens the case for vaccination in these birth cohorts and has implications for vaccination strategies, including the value and cost-effectiveness of enhanced formulations which have demonstrated superior immunogenicity and effectiveness in older adults^19^.

The US and Brazil hospitalization data indicate increased burden of severe disease due to A/H1N1pdm09, but whether this is due to changes in infection risk and/or changes in clinical severity warrants further investigation, particularly given that A/H1N1pdm09 was associated with relatively severe outcomes among hospitalized adults before the COVID-19 pandemic^20,21^. Our analyses rely on observational data and are susceptible to reporting biases; however, consistency across four surveillance systems with distinct ascertainment mechanisms and varying analysis methods makes a surveillance artifact implausible. Replication with additional age- and subtype-resolved surveillance data would further substantiate our findings.

Together, our results indicate that the earliest birth cohorts, who are the most clinically vulnerable and have historically been considered relatively protected from influenza A/H1N1pdm09, likely now face substantially elevated risk from these viruses. This carries potential implications for vaccine policy and healthcare capacity planning. Ensuring that surveillance infrastructure captures age- and subtype-resolved influenza data globally will be essential for tracking this shift and quantifying its consequences for the populations most at risk for severe influenza.

## Data availability

Brazilian hospitalisation data is available from https://dadosabertos.saude.gov.br/dataset/srag-2019-a-2026 and https://dadosabertos.saude.gov.br/dataset/srag-2013-2018. Australian counts are available from https://www.cdc.gov.au/resources/publications/nndss-dataset-influenza. Sequence metadata used in this analysis were retrieved from GISAID under identifier EPI_SET_260508ng (https://doi.org/10.55876/gis8.260508ng).

## Code availability

Code underlying analysis is available at https://github.com/AMC-LAEB/H1N1_age.

## Acknowledgements

We gratefully acknowledge all data contributors, i.e., the Authors and their Originating laboratories responsible for obtaining the specimens, and their Submitting laboratories for generating the genetic sequence and metadata and sharing via the GISAID Initiative, on which this research is based. We further acknowledge those responsible for the collection and curation of the NNDSS, SIVEP-Gripe, and FluSurv-NET surveillance datasets analyzed here.

## Competing interests

C.A.R. has received consulting fees from CSL Seqirus, Moderna, Pfizer, GSK, and Sanofi for advisory services unrelated to this work.

## Funding

S.P.J.d.J. and C.A.R. were supported by the Dutch Research Council (Nederlandse Organisatie voor Wetenschappelijk Onderzoek) Vici Award 09150182010027.

## Author contributions

Conceptualization: S.P.J.d.J., C.A.R.; Formal analysis: S.P.J.d.J.; Writing – original draft: S.P.J.d.J. – Writing – review & editing: S.P.J.d.J. & C.A.R.

## Online Methods

### Data

To investigate patterns of subtype-specific influenza burden by birth cohort, we assembled a set of publicly available, case-based surveillance datasets with subtype and age metadata spanning both the pre- and post-pandemic periods. Datasets were included if they provided data for at least two years both before and after 2020, age was reported with a resolution of maximum 5 year bins and age was not censored below 85 years of age (as censoring at younger ages would obscure patterns in the pre-1950 birth cohorts of primary interest). Three datasets meeting these criteria were identified and are described below.

The National Notifiable Diseases Surveillance System (NNDSS) dataset is publicly available through the Australian Government Department of Health and Aged Care and contains laboratory-confirmed influenza cases reported by states and territories across Australia. Confirmed cases are defined by isolation of influenza virus by culture from respiratory tract specimens, detection of influenza virus RNA by nucleic acid testing, detection of influenza A antigen, or serological evidence of seroconversion. Of 1,725,472 influenza records spanning 2 January 2015 to 3 January 2025, 93,850 were subtyped as A/H1N1pdm09 or A/H3N2 and were included in analyses. In this dataset, age is reported in five-year bins, with the oldest bin censored at 85 years of age.

The GISAID dataset consists of human-derived IAV isolates from GISAID (www.gisaid.org) with at least one sequenced gene segment, with collection dates between 2015 and 2025 and submission dates on or before 1 March 2026. Of 382,882 isolates meeting these criteria, 229,785 had associated host age metadata and were included in subsequent analyses.

Brazilian hospitalization data were drawn from SIVEP-Gripe (Sistema de Vigilância Epidemiológica da Gripe), Brazil’s influenza epidemiological surveillance system. This system captures cases of hospitalized severe acute respiratory infection (SARI), defined as influenza-like illness with at least one of dyspnea, respiratory distress, oxygen saturation below 95%, or cyanosis, as well as influenza deaths irrespective of hospitalization status. Datasets were obtained from the Open Data Portal of Brazil’s Unified Health System (Sistema Único de Saúde). Overall, the data includes 66,219 subtyped IAV records with associated birth year metadata (40,971 A/H1N1pdm09 and 25,248 A/H3N2), alongside 142,305 RSV and 10,961 HPIV records with birth year metadata, spanning 2015–2025.

Beyond these three case-based datasets, we extracted age- and subtype-specific influenza hospitalization data from a recent publication reporting US FluSurv-NET data (O’Halloran et al.^3^). FluSurv-NET is a population-based surveillance system that prospectively captures laboratory-confirmed influenza-associated hospitalizations across all age groups from more than 300 acute-care hospitals in the United States. From the published figures, we obtained age-specific subtype ratios, reflecting the cumulative hospitalization rate for A/H1N1pdm09 compared to A/H3N2 for each age bin, normalized such that this subtype ratio equals one among infants aged 0-11 months. We obtained these for each individual season 2015/2016 through 2019/2020 as well as 2023/2024 and 2024/2025. Estimates and 95% CIs were digitized from figures using WebPlotDigitizer v5.2 (https://automeris.io/wpd/) because the source data are not publicly available^3^.

### Subtype odds ratios

For each dataset, birth year was inferred from the associated age metadata. For the GISAID data, birth year was computed as the rounded decimal collection date minus the recorded age in years. For the Brazilian SIVEP-Gripe data, birth year was taken directly from the date of birth field where available. For the Australian NNDSS data, where age is reported in five-year bins, birth year was imputed as the collection year minus the midpoint of the reported age bin (lower bound plus two years), introducing a maximum misclassification of ±2 years. For the FluSurv-NET data, birth year was estimated within each season by subtracting the age from the season year.

To characterize birth cohort-specific differences in the relative odds of A/H1N1pdm09 versus A/H3N2 infection, cases were aggregated into birth year bins over the range 1930– 2000, with bin widths of 3 years for the GISAID, Brazilian, and US datasets and 5 years for the Australian NNDSS dataset. For each bin *i*, the subtype odds ratio was computed as the ratio of A/H1N1pdm09 to A/H3N2 counts in bin *i*, normalized by the overall ratio across all bins:

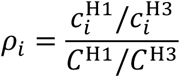

where 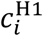 and 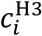 are the total A/H1N1pdm09 and A/H3N2 counts in bin *i*, and *C*^H1^and *C*^H3^ are the corresponding totals across all bins. Confidence intervals were derived on the log scale assuming Poisson counting error:

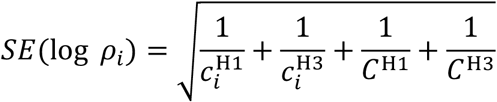

with 95% CIs obtained as exp (log *ρ*_*i*_ ± 1.96 ⋅ SE).

Ratios were computed separately for pre-pandemic (2015–2020) and post-pandemic (2023–2025) periods, with all available seasons pooled within each period. To place estimates on a common scale, ratios were normalized to the 1965–1974 reference birth cohort window by subtracting the median log ratio across bins whose midpoints fell within that window, computed separately within each period. For the Brazilian data, the analysis was repeated replacing either IAV subtype as the denominator with RSV or HPIV counts from the same dataset.

For the FluSurv-NET data, season-level estimates were pooled across seasons within the pre-pandemic (2015/2016 to 2019/2020) and post-pandemic (2023/2024 and 2024/2025) periods by inverse-variance weighting. As above, pooled ratios were normalized to the 1965–1974 reference birth cohort window by subtracting the median log ratio across bins whose midpoints fell within that window, within each period separately.

To estimate the timing of the shift in subtype odds ratios, we computed the A/H1N1pdm09 vs A/H3N2 subtype odds ratio comparing individuals born before 1950 to those born 1965–1974 in non-overlapping 6-month bins of GISAID sequence collection date, spanning 2015 to 2025. To control for compositional bias arising from over-represented countries, no single country was permitted to contribute more than 5% of sequences per bin; estimates were averaged across 10 subsampling replicates. Bins with fewer than 5 sequences for either subtype in either birth cohort were excluded. Odds ratios and 95% confidence intervals were computed on the log scale as described above for the subtype odds ratio analyses. Bins falling outside the defined pre-pandemic (2015–2020) and post-pandemic (2023–2025) periods are shown in grey in Fig. 1c.

### Relative risk of birth year

To assess whether the birth year distribution of sequenced A/H1N1pdm09 and A/H3N2 cases shifted between the pre- and post-pandemic periods we fitted separate negative binomial mixed-effects models for each subtype using GISAID sequence data, incorporating heterogeneity across countries. Unlike the subtype odds ratio analyses, this approach models the absolute birth year distribution of sequences for each subtype independently, allowing us to distinguish whether observed shifts in subtype odds ratios reflect increased A/H1N1pdm09 burden, decreased A/H3N2 burden, or both.

Sequences were aggregated into 2-year birth year bins within each country and period (pre-pandemic: 2015–2020; post-pandemic: 2023–2025), retaining only country-period combinations with at least 2 sequences per bin and countries contributing data in both periods.

To account for progressive depletion of older birth cohorts by mortality, the log of the cohort- and country-specific survival proportion (derived from UN World Population Prospects period life tables (from the wpp2024 R package) relative to a 2014 baseline, with a pooled cross-country average used as a fallback for countries lacking WPP data) was included as an offset, such that the model outcome reflects sequenced cases per surviving individual rather than raw counts.

We fitted a model of the form:

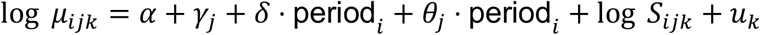

where *μ*_*ijk*_ is the expected sequence count for period *i*, birth year bin *j*, and country *k*; *γ*_*j*_ are birth year bin fixed effects; *δ* is the main effect of period (i.e., pre- vs post-pandemic); *θ*_*j*_ are bin-specific period interaction coefficients, with the full set of interaction terms allowing the birth year distribution to differ freely between periods; *S*_*ijk*_ is the survival proportion offset; and *u*_*k*_ ∼ *N*(0, *σ*^2^) is a country-level random intercept accounting for between-country heterogeneity in sequencing practice. All estimates were expressed relative to the 1965–1974 reference birth cohort window, with per-bin relative rates and 95% confidence intervals derived via the delta method. Models were fitted using glmmTMB version 1.1.14 in R version 4.5.1.

### Pooled estimates

To obtain a pooled estimate of the fold change in A/H1N1pdm09 burden by birth cohort, we combined estimates derived from seven independent analyses spanning four data sources: two analyses of GISAID sequence data (A/H1N1pdm09 vs A/H3N2 subtype odds ratios, and A/H1N1pdm09 birth year representation), three analyses of Brazilian hospitalization data (A/H1N1pdm09 vs A/H3N2, A/H1N1pdm09 vs RSV, and A/H1N1pdm09 vs HPIV), the subtype odds ratios in the Australian NNDSS surveillance data, and the subtype odds ratios in the US FluSurv-NET hospitalization data.

For each analysis, we calculated the birth cohort-specific fold change by taking the difference in normalized log odds ratios (relative to the 1965-1974 reference) between the post-pandemic (2023–2025) and pre-pandemic (2015–2020) periods within each birth year bin and exponentiating to obtain the fold change on the ratio scale. For the count odds ratio analyses (A/H1N1pdm09 vs A/H3N2, A/H1N1pdm09 vs RSV, and A/H1N1pdm09 vs HPIV), the log fold change was computed as the difference between the post- and pre-pandemic normalized log subtype odds ratios within each birth year bin, with standard errors derived as 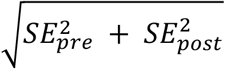. For the GISAID birth year enrichment analysis, the post vs pre contrast was computed from the mixed-effects model estimates via the delta method, combining the within-period contrasts on the log scale. For each analysis, the resulting log fold change and its standard error were expressed relative to the 1965–1974 reference birth cohort.

Because multiple estimates were drawn from the same underlying dataset (two from GISAID and three from Brazil), we used a multilevel random-effects pooled analysis implemented via the rma.mv function in the R package metafor, with individual analysis nested within data source. This structure accounts for the non-independence of estimates derived from the same underlying data. All models were fitted by restricted maximum likelihood (REML). Pooled estimates and 95% confidence intervals were obtained by exponentiating the model intercept and its standard error. Analyses were performed using metafor version 5.0.1 in R version 4.5.1.

To assess the robustness of pooled fold-change estimates to the choice of reference birth cohort, we repeated the pooled analysis using seven alternative reference windows spanning 1960–1999, each ten years wide and stepping in five-year increments (1960– 1969, 1965–1974, 1970–1979, 1975–1984, 1980–1989, 1985–1994, and 1990–1999). For each candidate reference window, all seven constituent estimates were re-normalized to that window prior to pooling, and the resulting fold-change estimates for each focal birth cohort bin from 1930 to 1995 were recorded (Extended Data Table 1). Pooled estimates were consistent across reference windows spanning 1960–1989, supporting the robustness of the primary analysis to the specific choice of reference cohort within this range. Estimates derived using reference windows anchored in the 1990s (1985– 1994 and 1990–1999) diverged somewhat from the others with higher fold increase estimates; however, this reflects the fact that pre- and post-pandemic subtype odds ratios are not as stable for approximately post-1990 birth cohorts (as visible in Fig. 1a). These reference windows are therefore less appropriate as a neutral baseline.

### Clade analysis

To assess the temporal emergence of the 6B.1A.5a.2a A/H1N1pdm09 clade in relation to the observed epidemiological shift, we computed the proportion of A/H1N1pdm09 sequences belonging to clade 6B.1A.5a.2a in non-overlapping 6-month bins of GISAID sequence collection date, spanning 2015 to 2025. Only sequences with an annotated clade designation were included. As above, no single country was permitted to contribute more than 5% of sequences per bin, with estimates averaged across 10 subsampling replicates, and bins with fewer than 5 sequences were excluded. Within each replicate and bin, the clade proportion was computed as the fraction of A/H1N1pdm09 sequences assigned to clade 6B.1A.5a.2a, with 95% confidence intervals derived from binomial sampling variance.

---

**Extended Data Fig. 1.**
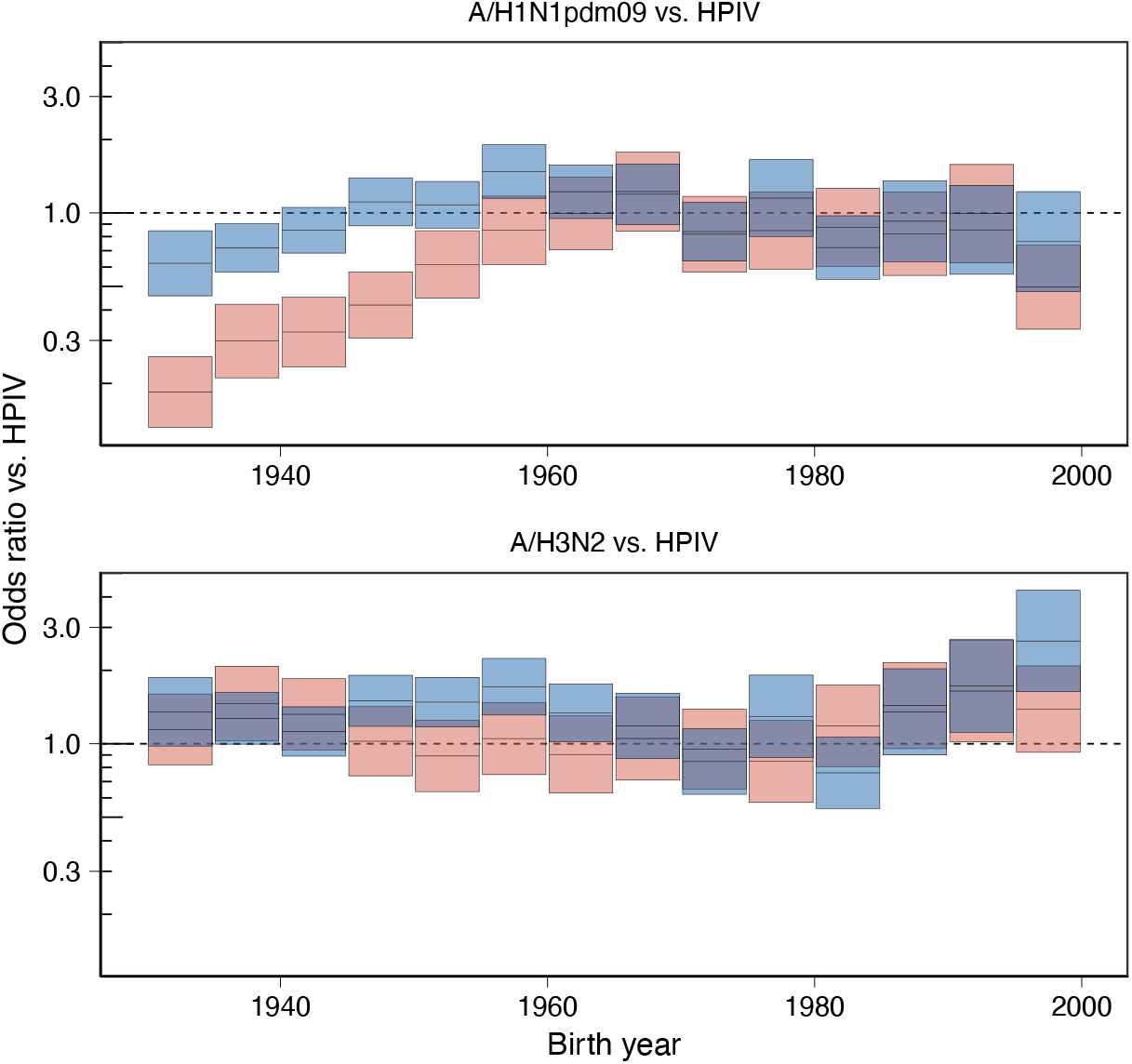
Odds a subtyped IAV is A/H1N1pdm09 (top) or A/H3N2 (bottom) relative to HPIV by birth cohort, relative to the 1965-1974 reference cohort in the Brazilian data, as in Fig. 1e

**Extended Data Fig. 2.**
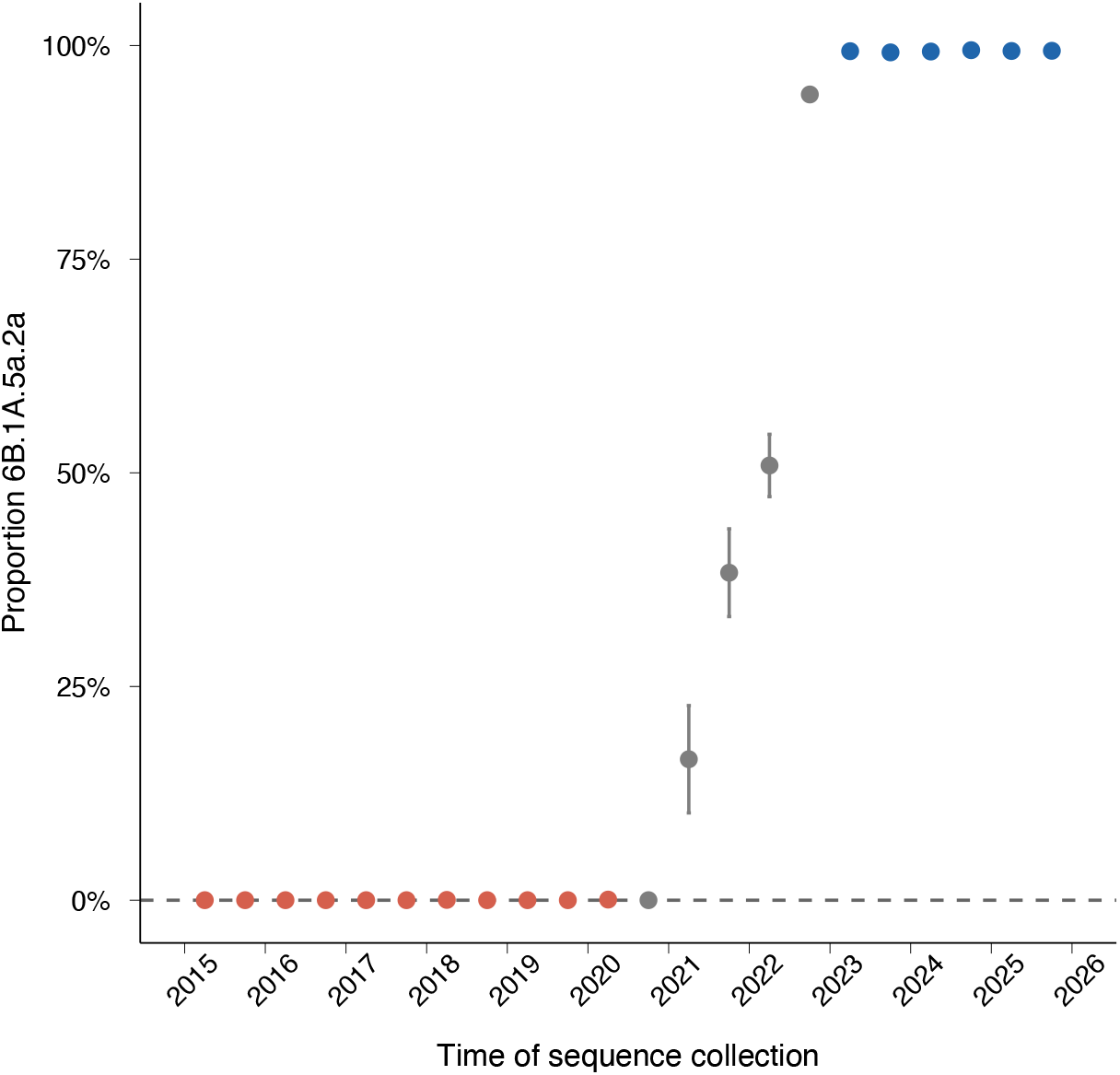
Proportion of A/H1N1pdm09 sequences belonging to clade 6B.1A.5a.2a in non-overlapping six-month bins from GISAID sequence data, spanning the pre-pandemic (2015–2020, red) and post-pandemic (2023–2025, blue) periods. Points represent bin midpoints and error bars show 95% confidence intervals based on binomial sampling variance. To control for compositional bias from over-represented countries, no single country contributed more than 10% of sequences per bin; estimates are averaged across 10 subsampling replicates. Grey points correspond to time bins outside the henceforth defined pre- (2015-2020) and post-pandemic (2023-2025) periods.

**Extended Data Table 1.** For each reference birth cohort (columns), rows show the estimated pooled fold increase in A/H1N1pdm09 by birth cohort and associated 95% CI. Italicized cells indicate those where the focal birth cohort is contained within the reference cohort.

|  | **1960-1969** | **1965-1974** | **1970-1979** | **1975-1984** | **1980-1989** | **1985-1994** | **1990-1999** |
| --- | --- | --- | --- | --- | --- | --- | --- |
| **1930-1934** | 3.12 (2.81-3.46) | 3.10 (2.58-3.72) | 2.91 (2.22-3.82) | 3.05 (2.28-4.08) | 3.01 (2.03-4.47) | 3.21 (2.21-4.66) | 3.37 (2.48-4.58) |
| **1935-1939** | 2.87 (2.59-3.18) | 2.82 (2.50-3.19) | 2.64 (2.24-3.10) | 2.70 (2.30-3.16) | 2.77 (2.25-3.41) | 2.91 (2.32-3.65) | 3.02 (2.38-3.82) |
| **1940-1944** | 2.66 (2.32-3.05) | 2.67 (2.38-3.00) | 2.59 (2.19-3.07) | 2.64 (2.25-3.09) | 2.67 (2.31-3.09) | 2.83 (2.37-3.37) | 3.03 (2.39-3.83) |
| **1945-1949** | 1.90 (1.80-2.01) | 1.95 (1.84-2.08) | 1.89 (1.70-2.11) | 1.91 (1.71-2.14) | 1.92 (1.67-2.21) | 2.06 (1.77-2.40) | 2.23 (1.88-2.64) |
| **1950-1954** | 1.43 (1.35-1.51) | 1.43 (1.26-1.61) | 1.43 (1.18-1.74) | 1.42 (1.19-1.69) | 1.42 (1.16-1.74) | 1.54 (1.24-1.91) | 1.67 (1.32-2.11) |
| **1955-1959** | 1.18 (1.00-1.40) | 1.22 (1.08-1.37) | 1.20 (1.10-1.31) | 1.21 (1.12-1.30) | 1.21 (1.03-1.41) | 1.30 (1.13-1.48) | 1.42 (1.28-1.58) |
| **1960-1964** | *1.01 (0.96-1.06)* | 1.04 (0.96-1.12) | 1.03 (0.88-1.19) | 1.02 (0.88-1.17) | 1.02 (0.86-1.21) | 1.10 (0.92-1.32) | 1.19 (0.98-1.45) |
| **1965-1969** | *0.98 (0.92-1.03)* | *1.00 (0.93-1.08)* | 1.00 (0.84-1.17) | 0.99 (0.82-1.18) | 0.98 (0.82-1.19) | 1.07 (0.87-1.30) | 1.15 (0.92-1.43) |
| **1970-1974** | 0.99 (0.85-1.16) | *1.00 (0.92-1.08)* | *0.99 (0.93-1.04)* | 0.99 (0.92-1.07) | 1.01 (0.95-1.07) | 1.11 (1.05-1.18) | 1.19 (1.07-1.34) |
| **1975-1979** | 0.96 (0.81-1.14) | 0.99 (0.90-1.10) | *1.00 (0.93-1.06)* | *0.98 (0.89-1.08)* | 1.00 (0.86-1.16) | 1.06 (0.92-1.23) | 1.19 (1.07-1.32) |
| **1980-1984** | 0.99 (0.83-1.17) | 1.00 (0.89-1.13) | 0.99 (0.87-1.12) | *1.01 (0.93-1.10)* | *1.03 (0.92-1.15)* | 1.09 (0.96-1.23) | 1.16 (0.97-1.38) |
| **1985-1989** | 0.99 (0.80-1.22) | 1.00 (0.88-1.14) | 1.01 (0.89-1.15) | 1.01 (0.87-1.18) | *1.01 (0.96-1.07)* | *1.10 (1.02-1.18)* | 1.20 (1.03-1.40) |
| **1990-1994** | 0.83 (0.69-1.01) | 0.83 (0.73-0.95) | 0.82 (0.74-0.92) | 0.82 (0.74-0.92) | 0.83 (0.74-0.94) | *0.90 (0.84-0.97)* | *1.00 (0.91-1.09)* |
| **1995-1999** | 0.84 (0.65-1.07) | 0.85 (0.72-1.01) | 0.81 (0.74-0.90) | 0.83 (0.74-0.94) | 0.86 (0.71-1.05) | 0.89 (0.75-1.05) | *0.99 (0.90-1.09)* |

